# Prospective genomic surveillance reveals cryptic MRSA outbreaks with local to international origins among NICU patients

**DOI:** 10.1101/2021.07.14.21260339

**Authors:** Jay N. Worley, Jessica W. Crothers, William J. Wolfgang, Sai Laxmi Gubbala Venkata, Maria Hoffmann, Victor Jayeola, Michael Klompas, Marc Allard, Lynn Bry

## Abstract

MRSA infections cause significant morbidity and mortality in neonates. Clinical testing and routine surveillance screening identified an increase in neonates with MRSA colonization and infection which triggered prospective genomic surveillance. Here we show the complex transmission dynamics of MRSA in a NICU setting. Analyses revealed concurrent transmission chains affecting 16 of 22 MRSA-colonized patients (68%), and 3.1% of all NICU patients (n=517). Prematurity and longer lengths of stay increased risks for colonization. Intervals of up to 7 months occurred among some cluster-related isolates. 3 of 22 MRSA-colonized patients developed invasive infections with the colonizing strain. Comparisons with 21,521 isolates in the NCBI Pathogen Detection Resource revealed NICU strains to be distinct from MRSA seen locally and internationally. Integration of international strain datasets in analyses increased the resolution of strain clusters and helped rule-out suspected transmission events. Analyses also identified sequence type 1535 isolates, emergent in the Middle East, carrying a unique SCC*mec* with *fusC* and *aac(6’)-Ie/aph(2’’)-1a* that provided a multi-drug resistant phenotype. NICU genomic surveillance identified cryptic MRSA colonization events, including NICU-endemic strains not linked with local hospital or international clusters, and has rich potential to guide improvements in infection prevention for this vulnerable patient population.

## Introduction

*Staphylococcus aureus* is a common cause of healthcare associated infections^1,2^. The pathogen can cause sepsis, respiratory and skin and soft tissue infections and is a major cause of patient morbidity and mortality^3^. Methicillin-resistant *Staphylococcus aureus* (MRSA) colonization is a well-recognized risk factor for infection^4,5^. Efforts to reduce infections have focused on decreasing MRSA transmission via institutional screening and decolonization^6,7^.

Neonatal populations in general, and pre-term and low birthweight infants in particular, are at increased risk of MRSA colonization and infection^8-11^ given their lack of protective colonizing microbiota and immature immune system^4,12,13^. Nosocomial transmission of *S. aureus* in neonates can occur among patients^14-17^, as well as from colonized healthcare workers^18-20^, parents^21,22^, and environmental reservoirs^23^. However, the relative contribution of these reservoirs remains controversial^13,24^. Additionally, neonatal intensive care units (NICUs) have witnessed a shift over the past twenty years from hospital acquired MRSA strains to community-acquired MRSA strains, requiring modifications in infection control practices for this patient population^3,25^.

Whole genome sequencing (WGS) enables epidemiologic outbreak investigations through high-resolution strain tracking^26-29^. While *S. aureus* outbreak investigations have historically used targeted molecular strain-typing, the discriminatory power of conventional techniques has been largely insufficient to establish definitive genomic and strain-specific linkages^30^. In contrast, WGS enables definitive clonal assessments and associated temporal relationships when combined with epidemiologic data^31^. Applied to NICU-populations, WGS has improved the detection of patient-to-patient^17,27^ and healthcare worker transmission events^32-34^, environmental reservoirs of infection^23,35^, and supported more effective MRSA screening programs^36^. Importantly, WGS can also rule-out nosocomial transmission events, providing reassurance to staff and appropriate utilization of hospital resources^29,37^.

The staphylococcal cassette chromosome *mec* (SCC*mec*) confers broad-spectrum resistance to beta-lactam antibiotics and is a common genetic cause for the MRSA phenotype. SCC*mec*s contain a *mec* gene encoding PBP2A, a peptidoglycan transpeptidase homolog of PBP2 with reduced affinity for beta-lactam antibiotics, particularly cephalosporins^38^. Many SCC*mec* types, as well as non-*mec* SCCs, encode other resistance genes such as the fusidic acid resistance gene *fusC* ^38,39^.

Observations of an increase in NICU MRSA colonizations and infections raised concerns for transmission events among neonates. Genomic analyses identified multiple cryptic outbreaks missed by clinical surveillance and diagnostic testing and also identified strain recurrences in distinct time periods that could not be attributed solely to patient-to-patient transmission. Expanded genomic analyses with >20,000 internationally deposited MRSA genomes in the NCBI Pathogen Detection resource significantly improved the phylogenetic resolution of local strain clusters and their dynamics of spread within the NICU, demonstrating the power of large-scale comparative genomic analyses for local hospital efforts to improve clinical and epidemiologic MRSA outbreak investigations.

## Results

### Antibiotic resistance phenotypes of NICU MRSA trigger prospective genomic surveillance

MRSA NICU isolates were evaluated phenotypically for similar antibiotic resistance profiles during a 341-day period over 2018-2019. Two suspected outbreaks during this targeted surveillance period triggered prospective genomic surveillance of all MRSA isolates from days 342-558, in addition to sequencing of prior available isolates from days 1-341 (Figure 1, Supplemental Figure 1). Over the 558 day period, 58 MRSA isolates, 55 screening and 3 diagnostic, were collected from 22 NICU patients. Of these patients, 16 neonates of 517 admitted during the prospective genomic surveillance period had MRSA isolates identified, representing 3.1% of NICU patients.

**Figure 1.**
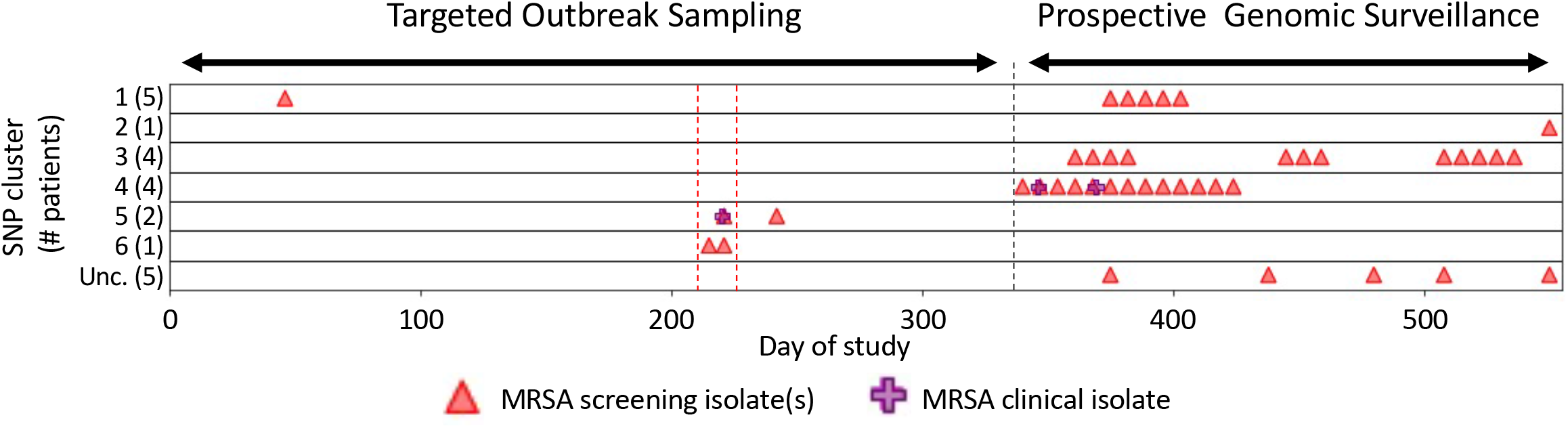
NICU outbreak cluster occurrence: X-axis indicates day during the analysis period. “Targeted Outbreak Sampling” refers to microbiologic-only assessments of NICU strains from days 0-341. Triangles and crosses indicate all occurring strains among patients that were also included in later genomic analyses. “Prospective surveillance” over days 342-558 indicates the period of prospective genomic analyses of new strains. Y-axis indicates NICU MRSA strains occurring in six NCBI, Pathogen Detection SNP clusters; numbers in parentheses indicate number of NICU strains from this study that occurred in the broader NCBI cluster, as follows: SNP Clusters (MLST, SCCmec type): **1**, PDS000067849.1 (8, IVa(2B)); **2**, PDS000069849.1 (72, IVa(2B); **3**, PDS000069850.1 (5, II(2A)); **4**, PDS000069851.1 (5, IVa(2B)); **5**, PDS000069882.1 (1535, unclassified SCCmec); **6**, PDS000069883.1 (8, IVa(2B)); Unc. = unclustered isolates with unrelated MLST and SCCmec types. SNP cluster 2 (NCBI outbreak cluster PDS000069849.1) contained only one isolate from this study (Supplemental File 2). Red dashed lines show a suspected outbreak during the, and a black dashed line marks when the Prospective Surveillance began.

### Prematurity and increased NICU lengths of stay elevate risks for MRSA colonization

MRSA colonization was significantly associated with prematurity as well as low-birth-weight compared to non-colonized patients (Table 1, q=0.002; Supplemental Table 1). In addition to prematurity and low birth weight, MRSA-colonized patients had longer average lengths of stay in the NICU than non-colonized patients at 52.5 days (±34.8 days) versus 20.0 days (±24.8 days) (p < 0.001). Prematurity was also associated with longer NICU lengths of stay than patients carried to term (defined as >36 weeks of gestation), at 29.7 days (±29.3 days) versus 6.8 days (±7.2 days), respectively (Supplemental Table 2).

### Genomic analyses identify 4 outbreak SNP clusters and unique NICU patient isolates

Genomic analyses using NCBI’s Pathogen Detection Resource identified 6 SNP clusters that accounted for 91.2% of sequenced NICU MRSA isolates originating from 17 of 22 MRSA-colonized patients (Figure 1). The remaining 5 patients with non-clustered MRSA strains each had short NICU admissions that yeilded only one isolate. These 5 patient-unique isolates showed no relationship to any *S. aureus* isolates deposited in NCBI and were not temporally linked to other NICU transmission events (Supplemental Figure 1).

SNP clusters 1, 3, 4, and 5, involved multiple patients (Figure 2). Clusters 1 and 3 were found across distinct time periods without patient overlap. The patient isolates in cluster 1 occurred in two separate intervals separated by 259 days, while strains in cluster 3 occurred across three time periods separated by 49 and 56 days (Figure 1). In contrast, clusters 4 and 5 occurred within single time periods. Among the two other NCBI-associated clusters, cluster 2 included one NICU isolate that clustered with an isolate reported from another institution, but was not clonally related (Supplemental File 2). Cluster 6 included two clonally-related isolates from the same patient that were separated by 5 SNPs. In total, 15 of 22 (68%) MRSA-colonized NICU patients, including all twins, and all cases of invasive disease occurred in clusters 1, 3, 4 and 5.

**Figure 2.**
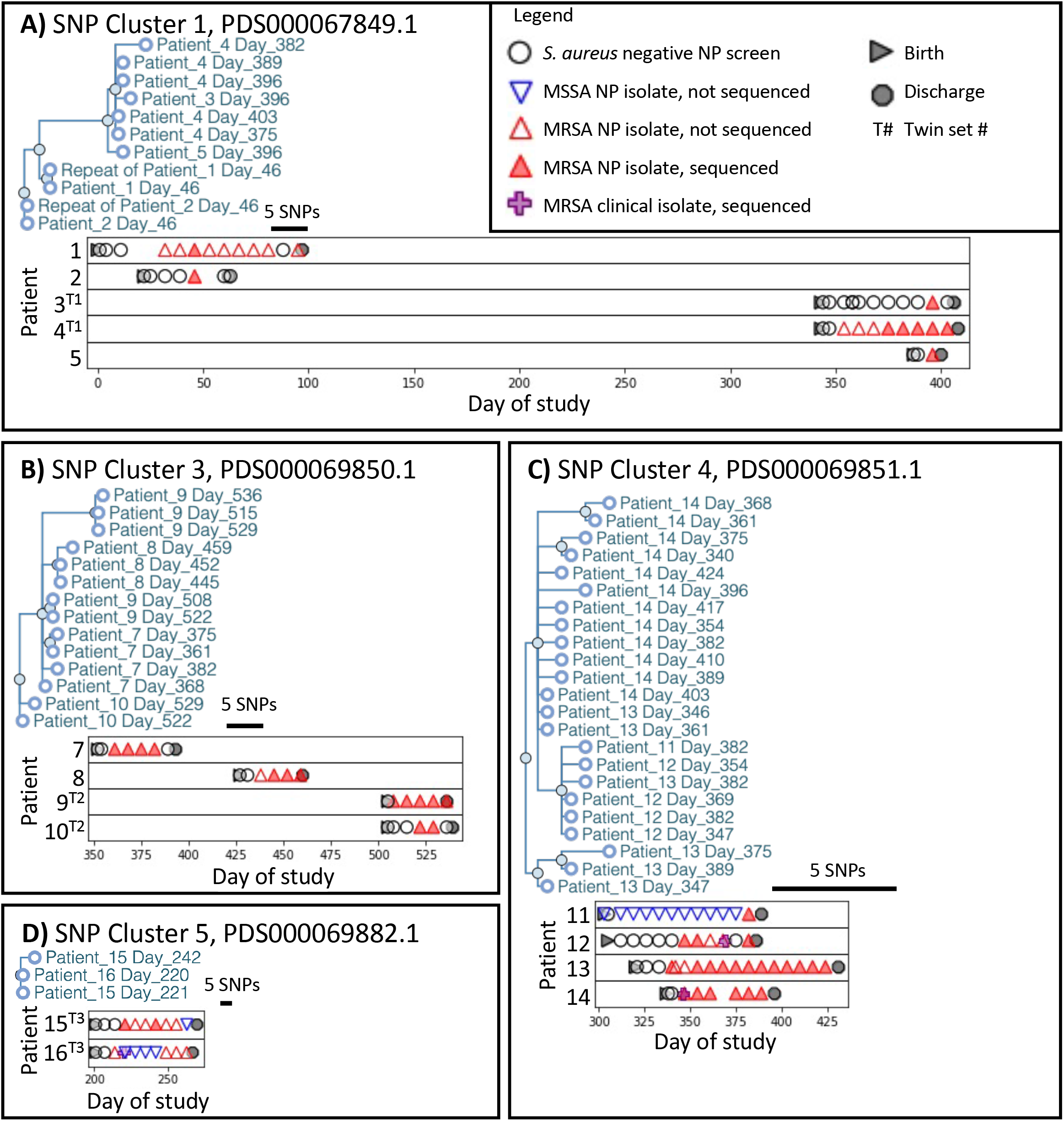
SNP cluster analyses reveal patient outbreaks and occurrence over time. Panels A-D show NICU MRSA strain phylogenetic trees from the NCBI Pathogen Genomes isolates browser with addition of the anonymized patientID and day of isolation. Graphs below each tree show the associated patients and strain occurrences relative to surveillance or diagnostic cultures. Though patient isolates fell within the broader NCBI SNP clusters, no other closely related strains from other institutions fell within these regions of the cluster SNP tree. Bar in each graph indicates 5 SNPs distance.

Comparisons of NICU strains in clusters 1, 3, 4 and 5 with adult nosocomial MRSA isolates seen at the hospital showed no clonal relationships between the populations, indicating that the outbreak clusters seen in the NICU were genomically distinct. Further, the NICU outbreak clusters did not include any of the 21,521 *S. aureus* isolate genomes deposited in NCBI’s Pathogen Isolates browser. To further assess nearest neighbor strains, whole-genome MLST (wgMLST) of NICU and 19,670 isolates with assemblies (Figure 3, Supplemental File 1, Supplemental File 2) confirmed that the NICU SNP clusters remained distinct even within broader wgMLST groups.

**Figure 3.**
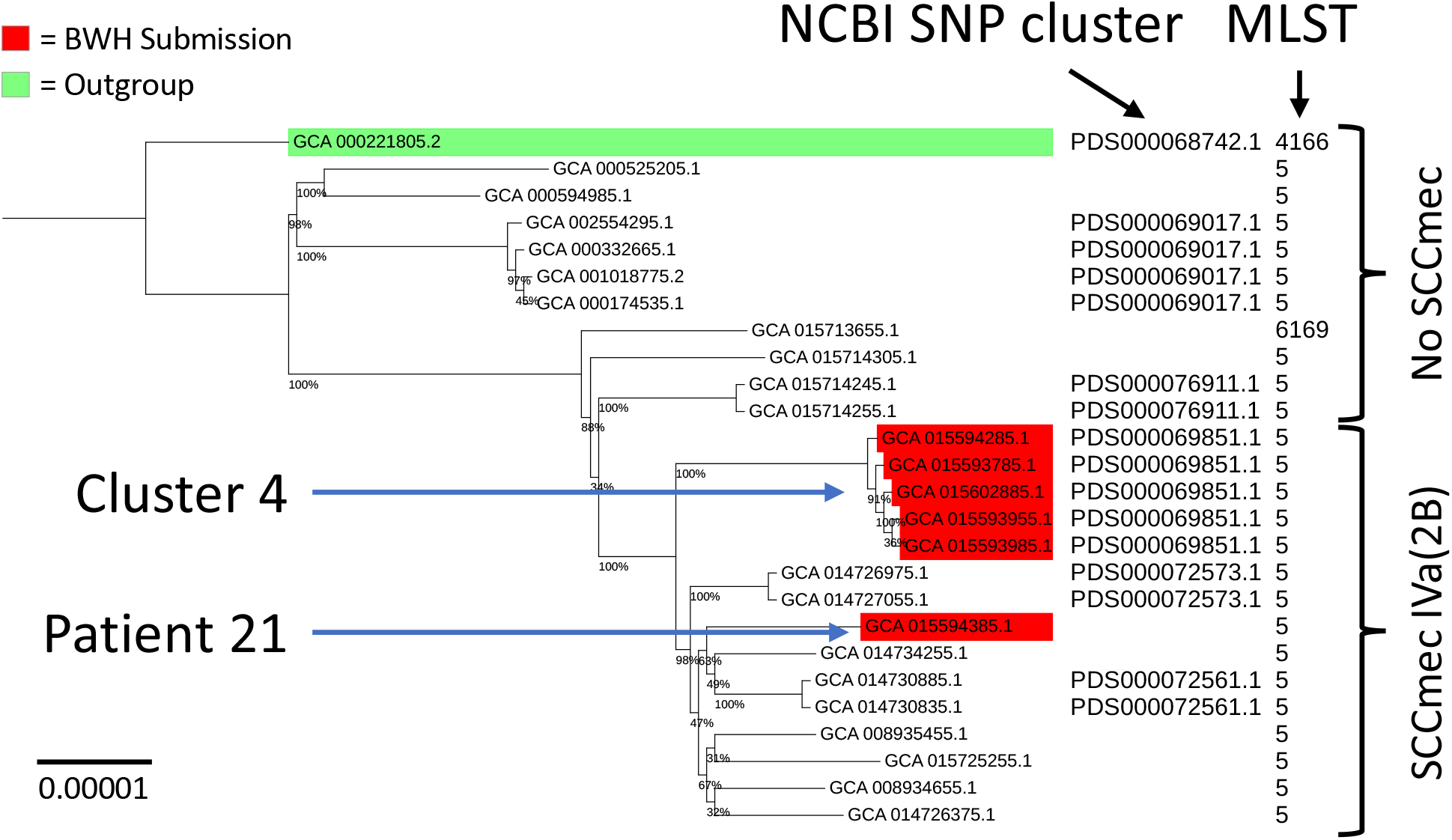
International MRSA strain datasets support effective rule out of local NICU suspected transmission events. Phylogenetic tree of NCBI ST5 SNP cluster isolates showing the NCBI SNP cluster, MLST and genomic SCCmec. The ST5 SNP clusters were downsampled to 5 representative isolates and wgMLST distances were used to define the 25 closest related strains in the NCBI Pathogen Detection program. SNPs within the wgMLST genes were used to calculate phylogenetic relationships Supplemental File 2). The outgroup strain (green) was most distantly related to the BWH strains by wgMLST SNPs. Bootstrap values are shown at phylogenetic nodes. NICU patient strains are shown in red, including 5 representative isolates from the 4 NICU patients in Cluster 4, and the additional MRSA isolate from patient 21. Bootstrap support values show placement of patient 21’s isolate in a separate sub-cluster distinct from Cluster 4.

### International MRSA genomic datasets enable rule-out of putative transmission events

Analyses of Patient 21’s MRSA isolate showed an identical antibiogram, MLST, and SCC*mec* type to isolates found in NICU cluster 4. However, incorporating the additional external strain genomic datasets showed clear placement of this patient’s isolate in a genomically distinct cluster (Figure 3, Supplemental File 1).

A gain of the SCC*mec* type IVa (2B) is evident in the sub-clade of multi-locus sequence type (MLST) 5 including this isolate. Eight of the nine other closely related SCC*mec* type IVa (2B) strains were submitted by Mount Sinai Hospital in New York City, New York, and the 9^th^ by the University of Pittsburgh Medical Center in Pittsburg, Pennsylvania. Thus, while Patient 21’s isolate and cluster 4’s isolates appear to belong to the same regional clade, this patient strain was ruled out as belonging to NICU cluster 4.

### Genomic identification of internationally-occurring MRSA in NICU patients

Cluster 5 isolates, from patients 15 and 16 in twin set 3, included two nasal-perirectal swab isolates from patient 15 and one epidermal abscess isolate from patient 16. The three isolates belonged to MLST 1535 (Figure 4A). The twin’s parents had recently immigrated from the Middle East where this MLST is more prevalent^40^. While ST1535 appears to be rare in the US, one ST1535 genome from a pediatric cystic fibrosis patient in Cincinnati (GCA_002124405.1) was included in analyses^41^. The MLST 1535 genomes share the presence of three antibiotic resistance genes; *aac(6’)-Ie/aph(2’’)-1a, fusC*, and *mecA* (Figure 4B). High-resolution closed genomes of the cluster 5 isolates revealed an SCC*mec* region containing these resistance genes flanked by multiple mobilizing insertion sequences and novel repetitive elements that had not been resolvable in the draft-level genomic data (Figure 4C). The SSC*mec* contains *ccrC* and *ccrAA* with *mecA* allele C2, *fusC* in the J3 region and an IS*256* flanked integron in the J1 region that contains *aac*(6’)-Ie-*aph*(2’’)-Ia^38^. Mobilizing IS*431*mec and IS*256* were each found multiple times in this region, as well as a set of three direct repeats sharing a 13 base long core sequence not found elsewhere in the chromosome. The cluster 5 isolates demonstrated fusidic acid resistance at 8 µg/ml, oxacillin resistance at 2 or 0.5 µg/ml, and gentamicin resistance at ≥16 µg/ml (Supplemental File 1).

**Figure 4.**
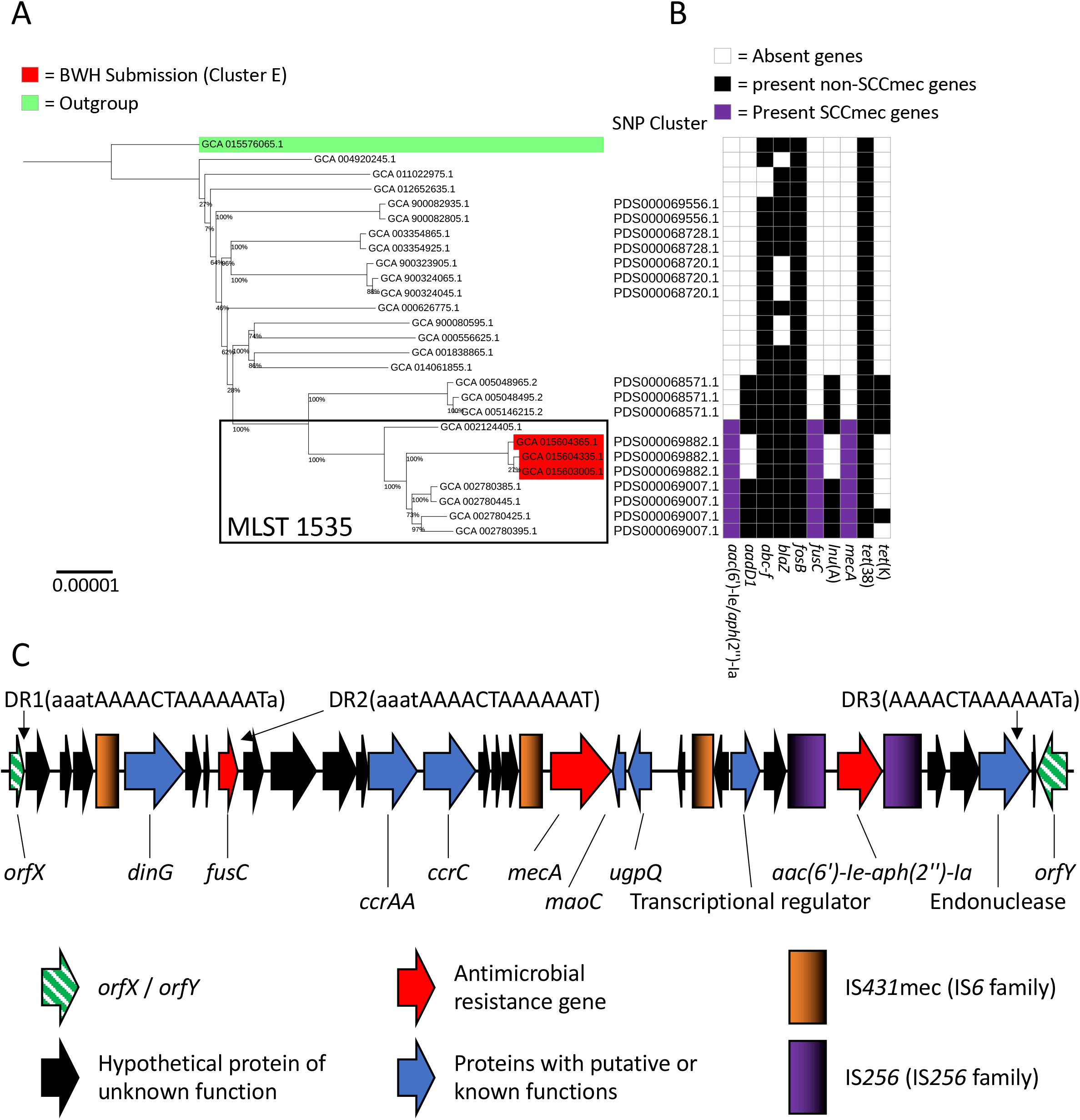
Genomic analyses identify NICU MLST 1535 strains with international clusters similar resistance gene profiles and variant SCC*mecs*. **A**. SNP based phylogeny of a wgMLST defined group of related strains reveals a clade of MLST 1535 strains from NICU Cluster 5 (red) that occurred in two patients. Strains were most similar to others identified in Saudi Arabia and one from Cincinnati. Strains outside of the MLST 1535 box are MLST 15. **B**. Antibiotic resistance gene profiles show the stepwise acquisition of resistance genes. The three NICU patient strains have lost *lnuA* and *aac(6’)-Ie/aph(2’’)-Ia*. **C**. A high-resolution genomic map of the SCC*mec* region from NCBI cluster 5 isolates closed by long-read genome sequencing. A 13 base AAACTAAAAAAT core sequence (DR1-DR3) with variable adjacent homologous sequence is directly repeated (DR) three times. This sequence does not appear elsewhere in the genome. Gradients in the IS element keys show directionality of the similar sequences.

## Discussion

Preterm neonates are highly susceptible to colonization with pathogens, including MRSA, per their lack of colonization with a complex microbiota, immature immune system and frequent co-morbidities associated with preterm birth. Genomic analyses of surveillance and diagnostic MRSA cultures from NICU patients identified multiple outbreak clusters and transmission events. Many of these transmissions were cryptic events that fell below thresholds of detection using standard phenotypic assessments with clinical microbiologic data.

Four MRSA outbreaks associated with patient-to-patient transmission, clusters 1, 3, 4 and 5, occurred during the period of prospective genomic surveillance (Figures 1 and 2). These clusters accounted for isolates in 15 of 22 MRSA-colonized patients (68%) and all invasive MRSA infections. Genomic analyses provided high resolution information to resolve clusters and to also exclude putative transmission events in cases where strains co-occurring in time had identical microbiologic phenotypes. Geomic analyses resolved co-occurring MRSA strains among patients into three distinct clusters 1, 3, and 4. In addition, the sequence type 5 (ST5) isolate seen in surveillance cultures from patient 21 showed identical microbiologic phenotypes to strains in cluster 4 but a genomically distinct SNP profile allowing it to be ruled out as part of a transmission chain (Supplemental File 1). The higher resolution analyses enabling this distinction used 21,520 publicly-available genomic datasets from NCBI to resolve sub-clusters among ST5 strains (Figure 3). The external MRSA genomic data greatly improved the phylogenetic resolution of locally obtained strains and was thus able to rule out strain relatedness and further transmission events. Incorporation of large genomic datasets from geographically diverse regions enhances the accuracy of local genomic investigations of nosocomial outbreaks^42^.

We identified two potentially endemic strains in the NICU. Clusters 1 and 3 occurred over discontinuous time periods (Figure 1). Cluster 1 involved a NICU strain seen prior to initiating prospective genomic surveillance, with only 9 SNPs difference between the point of first detection of the cluster and its recurrence 259 days later (Figure 2). This finding concurs with prior literature showing that persistent strain carriage or presence on surfaces may accumulate low levels of SNP heterogeneity over time^42-45^. Similarly, cluster 3 isolates showed no consistent SNP differences while also appearing in three discontinuous time periods separated by 56 days and 49 days. Both clusters raise the possibility of sustained reservoirs that can seed intermittent outbreaks.

Two of four patients with isolates in cluster 4 developed invasive disease. Patient 12 had positive MRSA surveillance cultures 3 weeks in advance of infection while patient 14’s infection was detected on the same day as positive surveillance cultures (Figure 2C). In all cases, longitudinal MRSA isolates from individual patients and among twin sets were clonally related. Surveillance cultures therefore have predictive value for patients at higher risk for infections. However, patients 11, 15 and 16 were also co-colonized by methicillin sensitive *S. aureus*, demonstrating that colonization by multiple *S. aureus* strains can occur in NICU patients and can shift over time.

The use of extensive and geographically diverse datasets improved NICU-focused assessments of strain reservoirs and potential transmission events. The ST1535 isolate seen in patients 15 and 16 from twin set 3 (Figure 4, Supplemental Table 1) occurred primarily in locations from the Middle East, including clinical and environmental reservoirs^46-48^. While the parents and visiting family had recent international travel from the Middle East, domestic sources cannot be ruled out, as evidenced by a similar strain found in Cincinnatti^41^. Recent observations of confirmed and suspected MDR SCCs encoding *mecA* with other antimicrobial resistance genes, including several with *fusC*, raises concerns globally for Staphylococcal infections requiring complicated antimicrobial therapy^49-51^. *fusC* fusidic acid resistance has been seen in non-*mec* SCC and SCC*mec* elements^49,52,53^. The SCC*mec* sequence from the closed isolate genomes were highly similar to draft SCC*mec* sequences reported by Senok, et al.^40^. Closing of these isolate genomes identified clear IS*431*mec and IS*256* sequences and additional direct repeats within the *SCCmec* that may facilitate accrual of additional resistance genes via further transposition or inter-recombination events with homologous mobile elements^54^.

All MRSA-colonized neonates had been admitted to the NICU for prematurity (Figure 2, Figure S1). Prematurity and associated long lengths of stay in the NICU elevate risks for MRSA colonization and infection^4,5^. Importantly, NICU-specific outbreak strains were found in 68% of MRSA-colonized patients (15 of 22 neonates). Prospective genomic surveillance thus offers a tool to rapidly identify related strains, aid the discovery of risk factors for transmission, define potential environmental or staff reservoirs, and thus inform interventions to halt further spread.

We demonstrate the application of longitudinal genomic surveillance in a NICU setting using international repositories of deposited strains to improve outbreak detection, including cryptic transmission events, and potential pathogen reservoirs. The addition of robust and gnomically diverse information improves the resolution of analyses and better enables critical rule-out determinations. Analyses also provide global context for uncommon strains that may be seen in an increasingly connected world.

## Methods

### IRB study protocol and data collection

The study was conducted under IRB protocol 2011-P-002883 (PI: Bry, Partners Healthcare). The Crimson LIMS was used for sample retrieval, and patient data and microbiological test results were retrieved using the Partners Research Patient Data Registry^55,56^. Data used for analyses were deidentified.

### Study design and clinical interventions

From days 1-341 MRSA isolates flagged from prior clinical microbiologic testing or nasal-perirectal swab screening were saved for phenotypic analyses of susceptibility profiles to evaluate suspicious outbreak clusters. Findings triggered a prospective genomic surveillance period from days 342 to 558 during which MRSA isolates from all nasal-perirectal swab cultures and diagnostic cultures were sequenced, in addition to the previously collected strains. Medical records for NICU patients during the surveillance period were evaluated for clinical diagnoses, reasons for NICU admission, and additional culture-based analyses. In the context of the clinical genomic surveillance program, only patient surveillance and diagnostic strains were evaluated. Non-clinical samples were not collected from patient family members or NICU personnel. Neonates colonized or infected with MRSA were placed in single rooms and managed with Contact Precautions.

### Sample collection

BD BBL CultureSwabs (BD, San Jose, CA) were used for nasal-perirectal (NP) screening for *S. aureus*. Swabs were cultured to colistin nalidixic acid (CNA) blood agar for 18-24 hours at 37°C. *S. aureus* colonies were identified by colony morphology, Gram stain and positive catalase and coagulase assays (Thermo). Diagnostic cultures with putative *S. aureus* colonies were evaluated similarly. One MRSA isolate from a patient that tested positive on a single nasal-perirectal screening swab was missed during the surveillance period and excluded from the study. Collected isolates were preserved at -80° C in 2ml Cryovials (Corning, Corning, NY). In the context of the clinical genomic surveillance program, and the IRB protocol under which it operates, only patient surveillance and diagnostic strains were evaluated. Non-clinical samples were not collected from patient family members or NICU personnel.

### Antibiotic resistance testing

*S. aureus* isolates were tested clinically for antibiotic resistance by VITEK 2 with Susceptibility Card AST-GP78 (Biomerieux, Durham, NC). Microdilution testing for fusidic acid resistance was done by broth microdilution according to CLSI guidelines^57^. MRSA isolates were called by resistance to cefoxitin.

### Genome sequencing

All isolate genomes were sequenced using Illumina NextSeq as described (Illumina, San Diengo, CA)^56^. Oxford nanopore sequencing (Oxford Nanopore, Cambridge, MA) and subsequent assembly using a combination of nanopore and Illumina reads was performed as previously described for MLST 1535 isolates^58^. Datasets were deposited under NCBI Bioproject PRJNA278886.

### Bioinformatic analyses

Patient data was visualized and analyzed using Python with the packages Matplotlib and SciPy^59,60^. Isolate genome sequencing reads were uploaded to the NCBI Sequence Read Archive for inclusion in the Pathogen Detection program (Supplemental table 1), which includes SNP clustering, virulence gene detection, antimicrobial resistance gene detection, and isolate metadata^61^. Phylogenetic results were visualized using the Pathogen Detection Isolates Browser.

Whole-genome MLST was performed using the program tblastn and coding sequences from RefSeq genome NC_007795.1^62,63^. The threshold for inclusion was an e-value ≤0.001 and a length between 0.5x and 1.5x the reference sequence coding length. Gene matches were retained if there was at most one high quality hit in each genome for all genomes in a downsampled set of all *S. aureus* isolate genomes available in the NCBI Pathogen Detection program, limited to 5 from each SNP cluster. Gene sequences were only kept if they matched the reference sequence length to allow for efficient SNP calculation across thousands of genomes. Genes were excluded if present in <90% of genomes, resulting in a database of 2021 genes. Genomes with <90% of these genes represented within the criteria above were excluded, resulting in a final database representing 15,477 genomes. From this set of genomic information, allelic differences were calculated to determine the closest set of 25 genomes to NICU MRSA isolates, keeping all tied for 25^th^ closest. SNP matrices were then created using the gene encoding sequences among these ≌25 genomes plus the NICU MRSA isolates. Phylogenies from these matrices were calculated with RAxML 8.2.11 using 1000 bootstraps and the GTR substitution matrix^64^.

Each strain’s MLST was calculated using the reference sequences and definitions from pubMLST.org^65^. SCC*mec* types were determined using the tools and guidelines from SCC*mec* Finder^66^. Virulence gene profiles and antibiotic resistance gene profiles were taken from the analysis at the NCBI Pathogen Detection Program ^61^.

Annotation of the MLST 1535 SCC*mec* region was performed as follows. First, the genome was annotated by the PATRIC RASTtk-enabled Genome Annotation Service^67^. Open reading frames within this sequence were confirmed or identified using blastp searches against the non-redundant protein sequences database at NCBI^63^. Cassette chromosome recombinase (*ccr*) gene identity was investigated using blastp and representative alleles using hits above 80% identity and length^40,68^.

## Supporting information

Supplemental File 1: Study Isolates

## Data Availability

MRSA strain genomic data are available under NCBI BioProject PRJNA278886.

https://www.ncbi.nlm.nih.gov/bioproject/?term=PRJNA278886

## Acknowledgements

The authors wish to thank Mary Delaney for clinical microbiologic support, the Wadsworth Center Advanced Genomic Technologies Center (AGTC) for sequencing, and Beth Flanigan, MD for helpful review of results. The work of JW was supported by the Intramural Research Program of the National Library of Medicine, National Institutes of Health, and the work of JC by the BWH Clinical Microbiology Fellowship. Studies were supported by P30 DK034854 (Bry). Sequencing at the AGTC was supported by FDA Cooperative Agreements 5U18FD006229 and 1U18FD006763 (Wolfgang).

## Notes

### Competing Interest Statement

The authors have declared no competing interest.

### Author Declarations

The study was conducted under IRB protocol 2011-P-002883 approved by the Massachusetts General-Brigham (MGB) IRB. The IRB protocol supports clinical operations in Infection Prevention and Clinical Microbiology. Patient microbiologic isolates and clinical metadata were de-identified for analyses.

