## Supplemental Figure 1 for "Prospective genomic surveillance reveals cryptic MRSA outbreaks with local to international origins among NICU patients"

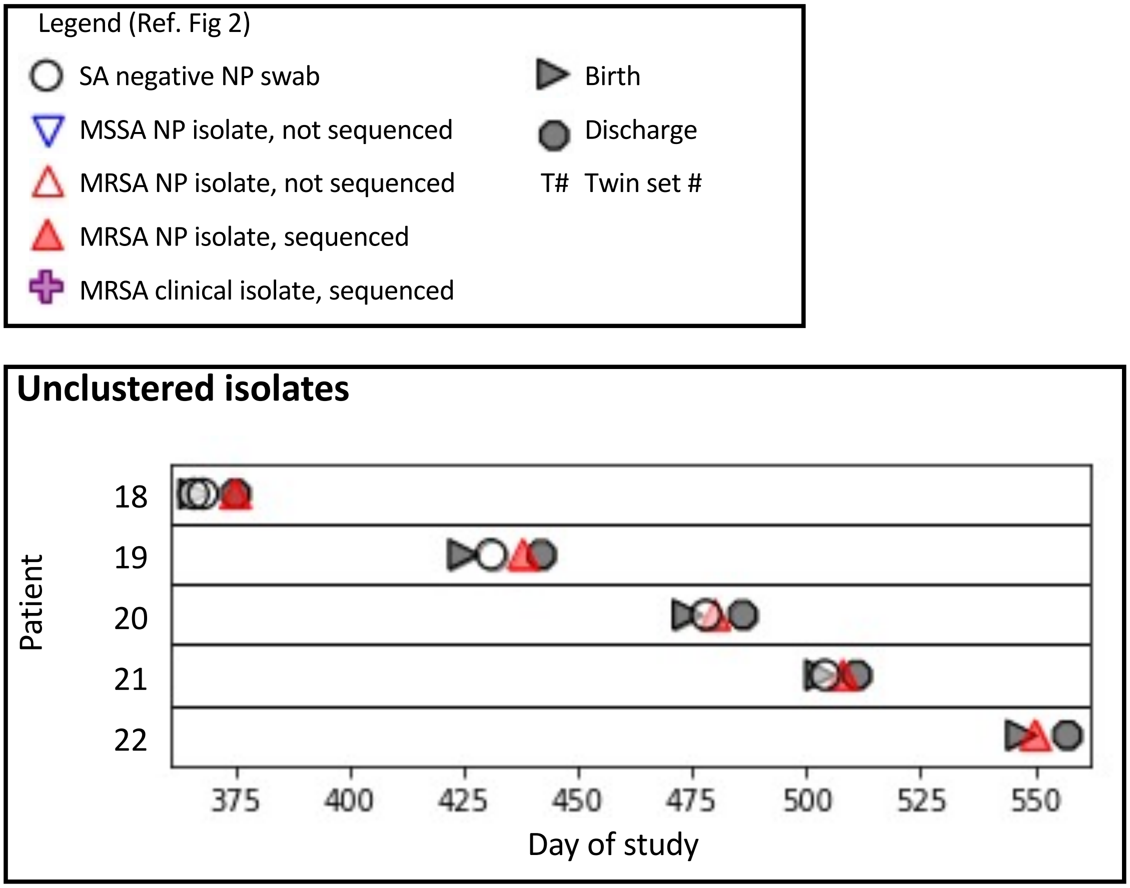


**Supplemental Figure 1. Unclustered patient isolates that were not linked to any in the NCBI Pathogen Detection program at the time of analyses**. MLST and SCCmec types are as follows: patient 18, MLST 8, SCCmec type IVg(2B); patient 19, MLST 8, SCCmec type IVa(2B); patient 20, MLST 87, SCCmec type IVa(2B); patient 21, MLST 5, SCCmec type IVa(2B); patient 22, MLST 6, SCCmec type IVa(2B). Refer to Figure 1 for the context of these isolates relative to those in other SNP clusters.
